# Kinematic Correlates of Early Speech Motor Changes in Cognitively Intact APOE-ε4 Carriers: A Preliminary Study Using a Color-Word Interference Task

**DOI:** 10.1101/2025.06.17.25329713

**Authors:** Mehrdad Dadgostar, Lindsay C. Hanford, Jordan R. Green, Brian D. Richburg, Averi Taylor Cannon, Nelson Barnett, David H. Salat, Steven E. Arnold, Marziye Eshghi

## Abstract

Alzheimer’s disease (AD) is the most prevalent form of dementia and a major public health challenge. In the absence of a cure, accurate and innovative early diagnostic methods are essential for proactive life and healthcare planning. Speech metrics have shown promising potential for identifying individuals with mild cognitive impairment (MCI) and AD, prompting investigation into whether speech motor features can detect elevated risk even prior to cognitive decline. This study examined whether speech kinematic features measured during a color-word interference task could distinguish cognitively normal APOE-ε4 carriers (E4+) from non-carriers (E4−). Lip movement properties were extracted across pre-, during-, and post-interference sentence segments. Descriptive statistics and independent t-tests were conducted to examine group-level trends in lip movement duration, average speed, and range. Although no group differences reached statistical significance, several features showed moderate effect sizes, suggesting potential neuromotor differences. A support vector machine model with a degree-2 polynomial kernel achieved 87.5% accuracy in classifying APOE-ε4 status using three features: lip movement duration prior to interference, average lip speed during interference, and the change in lip movement range from pre- to during-interference segments. These features reflect subtle differences between the two groups in baseline motor planning, susceptibility to cognitive-motor interference, and articulatory adaptability. The model’s high precision (88.90%), sensitivity (88.90%), and specificity (85.70%) underscore the potential clinical utility of speech kinematics for early risk identification. These findings support the use of non-invasive, low-burden speech analysis as a promising digital biomarker for AD risk in cognitively intact individuals and highlight its potential for scalable, remote screening and longitudinal monitoring in diverse populations.

## Introduction

Apolipoprotein E4 (APOE-ε4) is the major genetic risk factor for Alzheimer’s Disease (AD), its presence significantly increasing the likelihood of developing the condition.^1^ The presence of one APOE-ε4 allele raises the risk of AD by three to four times, while homozygous carriers face a 12- to 15-fold increase.^2–5^ Moreover, the APOE-ε4 allele exerts a profound influence on cognitive processes and motor control, particularly in the context of aging and neurodegenerative diseases.^6–9^

APOE-ε4 status has been associated with accelerated decline in memory, executive function, processing speed, attention, and spatial cognition.^7,10–12^ These deficits are linked to structural and functional brain changes, including reduced hippocampal volume, disrupted brain connectivity, and increased vulnerability to neurodegenerative processes.^13–17^ The allele’s role in mild cognitive impairment (MCI) further underscores its clinical relevance. APOE-ε4 status can increase susceptibility to MCI, particularly the amnestic subtype, and is associated with faster progression from MCI to AD.^18–21^ In addition, APOE-ε4 carriers experience more pronounced decline in motor abilities compared to non-carriers, which can manifest as decreased muscle strength, impaired coordination, and a greater risk of falls.^6,22–25^ The effects on motor function are attributed to apolipoprotein’s role in neuroplasticity, neuromuscular junction integrity, and overall brain health.^26–29^ Gait speed, an important marker of physical function, declines more rapidly in APOE-ε4 carriers, emphasizing the allele’s contribution to physical frailty in aging populations.^8,23,25^

### Cognitive-Motor Interactions in AD: Insights from Verbal Stroop Tasks

The *Stroop* task, a widely used neuropsychological test, effectively assesses cognitive control, particularly selective attention and inhibitory control.^30,31^ The task requires individuals to suppress automatic responses, such as reading a word, in favor of identifying the ink color in which the word is printed, creating a conflict between the two responses. Individuals with MCI and AD consistently perform worse on Stroop tasks compared to cognitively healthy older adults,^32–37^ exhibiting longer reaction times and greater error rates, particularly on incongruent trials where word-color and semantic meaning differ.^33,38^ These deficits are thought to reflect impairments in executive functions, including reduced inhibitory control and slower processing speed, both hallmark cognitive deficits in MCI and AD. Furthermore, Stroop task performance has been shown to correlate with underlying AD biomarkers, such as increased amyloid-β and tau pathology, making it a valuable tool for early detection.^39–41^

Variants of the Stroop task, including the oral Stroop task, introduce a verbal component, making it particularly relevant for studying speech motor control under cognitive load. In individuals with MCI and AD, the combination of cognitive conflict and the motor demands of speech can exacerbate performance deficits, leading to slower response times, increased speech hesitations, and more frequent articulation errors.^33,42^ These findings suggest that Stroop-inspired tasks can effectively probe the cognitive-motor interface, particularly in populations at risk for neurodegenerative diseases.

Speech alterations have long been recognized as a hallmark of AD, with prior studies linking AD-related neuropathology to changes in prosody, fluency, acoustics, and linguistic skills.^43–46^ These disruptions have made speech an attractive target for early diagnosis, particularly through acoustic analyses and self-reported communication difficulties. However, existing diagnostic approaches often rely on global acoustic features or subjective reports, which may lack the resolution needed to detect subtle neuromotor disruptions that precede overt cognitive decline. The long-term goal of this research is to advance speech as a sensitive and scalable biomarker for preclinical AD with the capability to capture early, subtle motor signatures before clinical symptoms emerge. Building on this motivation, the current study introduces the application of speech kinematic analysis to investigate cognitive-motor interactions in individuals genetically predisposed to AD. The primary aim of this study is to determine whether lip kinematic features recorded under minimal cognitive load can accurately classify individuals based on APOE-ε4 status (ε4+ vs. ε4−). By focusing on subtle motor signatures during low-demand speech tasks, this approach seeks to identify early, preclinical markers of AD risk that are scalable and non-invasive. We hypothesized that individuals with APOE-ε4 status would demonstrate early disruptions in speech motor performance compared to noncarriers.

Prior work from our group demonstrated that cognitively intact APOE-ε4 carriers exhibit early neuromuscular alterations in orofacial speech production, detectable via surface electromyography (EMG). While EMG effectively captures muscle activation patterns, it lacks the spatial resolution to fully characterize articulatory trajectories and inter-articulator coordination. Kinematic analysis, by contrast, enables precise quantification of the spatial and temporal dynamics of articulator motion. This approach has already shown diagnostic utility in detecting early bulbar dysfunction in neurodegenerative diseases such as amyotrophic lateral sclerosis (ALS),^47–50^ where kinematic markers have demonstrated greater sensitivity to early neuromotor decline than either acoustic measures or subjective reporting.

To probe these dynamics in the context of preclinical AD, we employed a modified color-word interference speech task designed to introduce mild cognitive conflict while preserving naturalistic speech production. Unlike conventional Stroop paradigms that rely on isolated words or short utterances, our task embeds cognitive interference within full sentences, maintaining linguistic continuity while subtly taxing cognitive control. This dual approach (i.e., leveraging the fine-grained sensitivity of lip kinematics and introducing controlled cognitive load) was designed to reveal early neuromotor vulnerability in ε4 carriers who remain clinically asymptomatic. Together, these innovations aim to overcome the limitations of traditional speech-based assessments and move the field toward more precise, high-resolution biomarkers of early AD risk. This design allows for the assessment of articulatory coordination as participants navigate mild cognitive conflict, offering a more ecologically valid and targeted window into the interplay between cognitive control and speech motor function.

## Methods

### Participants

The participants consisted of 16 older adults ranging in age from 50-90 years. Data was collected at the Speech Physiology and Neurobiology of Aging and Dementia (SPaN-AD) Lab as part of an ongoing longitudinal study. All participants in the study held an education level of at least 16 years. All participants were native speakers of American English and exhibited normal cognition as assessed by a neuropsychologist. The cognitive status of the participants was examined by a comprehensive array of standard neuropsychological tests that assess attention, processing speed, learning, memory, language, and executive function. Details of the neuropsychological assessments are provided by Eshghi et al.^51^ In addition, all participants scored 0 on the Clinical Dementia Rating (CDR) scale global score and less than 11 on the Geriatric Depression Scale (GDS). All participants exhibited normal, age-appropriate hearing status in at least one ear as verified by a pure tone audiometer (Earscan 3, Micro Audiometric Corp.) and normal visual acuity.^52^ None of the participants showed a clinical diagnosis of MCI, any neurological condition, or were using any psychoactive drugs. Participants were genotyped with respect to the APOE-*ε*4 gene and were put into two groups: (1) carriers of APOE-*ε*4 who possess heterozygous *ε*3-*ε*4 alleles or homozygous *ε*4-*ε*4 alleles and (2) noncarriers of APOE-*ε*4 who possess homozygous *ε*3-*ε*3 alleles. Characteristics of the participants in each group are provided in Table 1. The study was reviewed and approved by the MGB Institutional Review Board.

**Table 1.** Baseline Characteristics Stratified by Genotype Carrier Status.

| Variables | All participants | ε4<sup>-</sup> | ε4<sup>+</sup> | *p*-value |
| --- | --- | --- | --- | --- |
|  | (*n* = 16) | (*n* = 7) | (*n* = 9) |  |
| Female, *n* (%) | 8 (50.00) | 2 (28.57) | 6 (66.67) | 0.057<sup>†</sup> |
| Family history of AD, *n* (%) | 6 (37.50) | 2 (28.57) | 4 (44.44) | 0.515<sup>‡</sup> |
| Age, mean (*SD*) | 67.44 (7.98) | 69.86 (9.91) | 65.56 (6.06) | 0.337<sup>‡</sup> |
| Total years of education, mean (*SD*) | 15.81 (1.97) | 15.86 (1.68) | 15.78 (2.28) | 0.937<sup>‡</sup> |
| MMSE, mean (*SD*) | 29.50 (0.89) | 29.57 (1.13) | 29.44 (0.73) | 0.801<sup>‡</sup> |
| MoCA, mean (*SD*) | 27.81 (2.14) | 28.43 (1.51) | 27.33 (2.50) | 0.297<sup>‡</sup> |

MMSE = mini-mental state examination; MoCA = Montreal cognitive assessment; SD = standard deviation. <sup>†</sup> Chi-square test. <sup>‡</sup> t-test.

### Procedure

Lower lip movements were recorded during sentence production using three-dimensional electromagnetic articulography (EMA) (Wave; Northern Digital, Inc.). The EMA system tracked a small (2 mm) electromagnetic sensor affixed to the midline of the lower lip’s vermilion border, using a sampling rate of 100 samples per second. Participants performed color-word interference speech tasks while seated in a supported chair. Stimuli were displayed on a wall-mounted screen positioned approximately six feet in front of them. Audio recordings were obtained using a Williams Sound MIC094 head-mounted microphone positioned approximately 5 cm from the left corner of the lips. The signal was routed through a Focusrite Scarlett 2i2 audio interface and digitized at a sampling rate of 10,000 Hz. Audio and EMA kinematic data were recorded synchronously to ensure precise temporal alignment. Participants were instructed to read sentences at their natural speaking rate and loudness.

### Stimuli

The speech samples consisted of a block of four sentences containing color words, with the font color of each word being incongruent with its semantic meaning (see Table 2). All sentences had comparable syntactic structures, each consisting of 14 words and 18 syllables. To align with the study’s focus on lip movement properties, the sentences contained a high proportion of bilabial consonants.

**Table 2.** Speech stimuli used in the study.

| Color-Word Interference Condition |  |  |
| --- | --- | --- |
| Pre-Interference | During-Interference | Post-Interference |
| Pammy and Bobby picked | red, teal, brown, blue, and pink | poppies with their mommy. |
| Piper and Mimi bought | brown, pink, green, teal, and red | backpacks from my papa. |
| Bebe and Pippa pitched | green, blue, teal, pink, and brown | beanbags to the batboy. |
| Barbie and Mabel made | blue, red, pink, teal, and brown | bloomers for a baby. |

Participants were presented with eight blocks of color-word interference sentences. Within each block, the same four sentences were presented in a randomized order. The font color for each color word was also randomly changed across blocks. However, the font color never matched the semantic meaning of the written word in any of the sentences. Two of the four sentences were designated for analysis, while the remaining two served as foil sentences. Data from these foil sentences were collected but not included in the final analyses. Foil sentences were included to reduce predictability and prevent participants from anticipating target sentences, thereby promoting natural speech patterns and improving the reliability of kinematic and acoustic measurements. The speech stimuli were adapted from MacPherson,^53^ with adjustments made to align with the objectives of the current study.

Participants were instructed to read each sentence while disregarding the conflicting font color, requiring them to suppress the influence of incongruent visual information. This aspect of the task imposed a heightened cognitive load compared to non-interfered parts of the sentences. The increased cognitive demand specifically engaged key cognitive domains, including inhibition (overcoming the automatic tendency to process the font color), selective attention (focusing on the written word while filtering out irrelevant visual distractions), and working memory (maintaining task rules and resolving conflicts between competing stimuli), all of which are susceptible to impairment in MCI and AD ^54,55^.

### Kinematic Data

SMASH software^56^ was used to extract lip kinematic features from the three-dimensional lip movement time series recorded during the production of the entire target sentences. Lip movement duration (s), lip average speed of movement (mm/s), and lip range of movement (mm^3^) were calculated from three distinct sentence segments: the segment preceding the color-interference task (Pre-Interference), the segment corresponding to the color-interference task (During-Interference), and the segment following the color-interference task (Post-Interference). By assessing kinematic measures of lip movements during these segments, we aimed to gain insights into the anticipatory effects (Pre-task), direct effects (During task), and subsequent effects (Post-task) of the heightened cognitive load imposed on the motor system.

Additionally, difference measures, including Δ-movement duration, Δ-average speed, and Δ-range of movement, were calculated to assess the impact of the interference task. Difference measures were computed by subtracting the kinematic values of the During-Interference segment (i.e., the segment containing color interference) from those of the Pre- and Post-Interference segments. These measures are hereafter referred to as Δ_During-Pre_ and Δ_During-Post_. These measures provide valuable insights into how individual speakers adjust their motor planning and execution in response to the varying cognitive demands of each sentence segment.

### Statistical Analyses

Descriptive statistics were computed for each speech kinematic feature within sentence segments, and independent t-tests were performed to examine group differences. To support interpretation, Cohen’s d effect sizes were calculated to quantify the magnitude and direction of group differences. Although the primary goal of this study was to classify APOE-ε4 carriers (ε4+) and non-carriers (ε4−) using machine learning models trained on speech kinematic features, we also report descriptive and inferential statistics to provide additional interpretability and contextual insight into group-level trends. Given the limited sample size, our primary emphasis remains on the machine learning results for classifying APOE-ε4 carrier status based on kinematic speech features. All statistical analyses were performed in MATLAB (Version 24.2, R2024b; The MathWorks Inc., Natick, MA, USA).

### Classification

The purpose of this study was to identify lip kinematic features recorded during color-word interference speech tasks that can accurately classify APOE-ε4+ and APOE-ε4− individuals. Three sets of difference features were extracted to capture the dynamic characteristics of lip movement across task conditions. Specifically, lip movement duration, average speed of movement, and range of movement features were calculated 1) across the pre-, during-, and post-interference segments of the color-interference task, 2) on difference values from segments preceding the interference task (Δ_During-Pre_), and 3) on difference values from segments following the interference task (Δ_During-Post_), yielding a total of 15 features. These 15 features are listed in Table 4. All possible permutations of these fifteen features (n=32,767) were systematically evaluated based on classification accuracy (ACC), and the optimal set with highest ACC value was selected to ensure a robust and discriminative representation of the task.

**Table 3.** Mean (SD) values for each feature in E4+ and E4-groups.

| **Genotype: E4+** |  |  |  |
| --- | --- | --- | --- |
| **Feature** | **Pre-Interference** | **Dur-Interference** | **Post-Interference** |
| **Lip Movement Duration (s)** | 1.341 (0.147) | 2.156 (0.167) | 1.420 (0.157) |
| **Lip Average Speed of Movement (mm/s)** | 50.014 (14.850) | 33.93 (6.613) | 47.536 (12.224) |
| **Lip Range of Movement (mm<sup>3</sup>)** | 11.600 (1.939) | 11.396 (1.509) | 11.210 (2.149) |
| **Genotype: E4-** |  |  |  |
|  | **Pre-Interference** | **Dur-Interference** | **Post-Interference** |
| **Lip Movement Duration (s)** | 1.435 (0.184) | 2.274 (0.607) | 1.503 (0.167) |
| **Lip Average Speed of Movement (mm/s)** | 48.618 (11.292) | 35.959 (14.247) | 46.075 (10.903) |
| **Lip Range of Movement (mm<sup>3</sup>)** | 10.325 (1.481) | 11.110 (3.798) | 11.569 (3.109) |
| **Genotype: E4+** |  |  |  |
| | ΔDuring-Pre | ΔDuring-Post | |
| **Lip Movement Duration (s)** | 0.821 (0.130) | 0.799 (0.190) |  |
| **Lip Average Speed of Movement (mm/s)** | -16.084 (12.956) | -13.606 (12.341) |  |
| **Lip Range of Movement (mm<sup>3</sup>)** | -0.205 (1.119) | 0.186 (1.251) |  |
| **Genotype: E4-** |  |  |  |
| | ΔDuring-Pre | ΔDuring-Post | |
| **Lip Movement Duration (s)** | 0.839 (0.546) | 0.771 (0.566) |  |
| **Lip Average Speed of Movement (mm/s)** | -12.659 (14.464) | -10.116 (8.045) |  |
| **Lip Range of Movement (mm<sup>3</sup>)** | 0.609 (2.784) | -0.459 (1.112) |  |

**Table 4.** Statistical comparison of lip kinematic features between E4+ and E4− groups with effect sizes and 95% confidence intervals.

| Feature | t-test<br>(p-value) | Effect Size<br>(Cohen's d) | Confidence Intervals |  |
| --- | --- | --- | --- | --- |
|  |  |  | Lower | Upper |
| Lip Movement Duration (s): Pre-Interference | 0.308 | -0.534 | -1.527 | 0.480 |
| Lip Movement Duration (s): Dur-Interference | 0.635 | -0.248 | -1.229 | 0.742 |
| Lip Movement Duration (s): Post-Interference | 0.333 | -0.484 | -1.426 | 0.475 |
| Lip Average Speed of Movement (mm/s): Pre-Interference | 0.840 | 0.099 | -0.859 | 1.052 |
| Lip Average Speed of Movement (mm/s): Dur-Interference | 0.738 | -0.176 | -1.130 | 0.784 |
| Lip Average Speed of Movement (mm/s): Post-Interference | 0.811 | 0.118 | -0.840 | 1.071 |
| Lip Range of Movement (mm3): Pre-Interference | 0.230 | 0.601 | -0.392 | 1.571 |
| Lip Range of Movement (mm3): Dur-Interference | 0.857 | 0.096 | -0.861 | 1.049 |
| Lip Range of Movement (mm3): Post-Interference | 0.802 | -0.128 | -1.082 | 0.830 |
| ΔDur-Pre of Lip Movement Duration (s) | 0.935 | -0.041 | -1.054 | 0.974 |
| ΔDur-Pre of Lip Average Speed of Movement (mm/s) | 0.640 | -0.236 | -1.190 | 0.727 |
| ΔDur-Pre of Lip Range of Movement (mm3) | 0.490 | -0.371 | -1.329 | 0.600 |
| ΔDur-Post of Lip Movement Duration (s) | 0.905 | 0.062 | -0.920 | 1.042 |
| ΔDur-Post of Lip Average Speed of Movement (mm/s) | 0.524 | -0.310 | -1.266 | 0.657 |
| ΔDur-Post of Lip Range of Movement (mm3) | 0.309 | 0.511 | -0.473 | 1.476 |

Classification of E4+ versus E4− was performed using a Support Vector Machine (SVM) classifier with a degree-2 polynomial kernel. Model performance was evaluated using the leave-one-out cross-validation method. Classification metrics, including accuracy, sensitivity, precision, and specificity, were calculated and reported to comprehensively assess the model’s discriminative capability.

## Results

Descriptive statistics for each lip kinematic feature across sentence segments are presented in Table 3. Group comparisons between ε4+ and ε4− individuals are summarized in Table 4, including independent t-test p-values, Cohen’s d effect sizes, and 95% confidence intervals. While none of the group differences reached statistical significance, likely due to the limited sample size, several features exhibited moderate effect sizes, suggesting potential trends that warrant further investigation in larger samples.

The classification accuracy of lip movement features during the color-word interference speech task was evaluated across two genetically distinct groups (E4+ and E4−). We used an SVM with a degree-2 polynomial kernel and leave-one-out cross-validation as a classification model. Among the 15 extracted parameters, the combination of lip movement duration during the pre-interference segment, average movement speed during the interference segment, and the ΔDuring–Pre difference in movement range yielded the highest classification accuracy. The best classification performance demonstrated an accuracy of 87.50%, with additional performance metrics including a Precision of 88.90%, a Sensitivity of 88.90%, and a Specificity of 85.70% (Figure 1).

**Figure 1.**
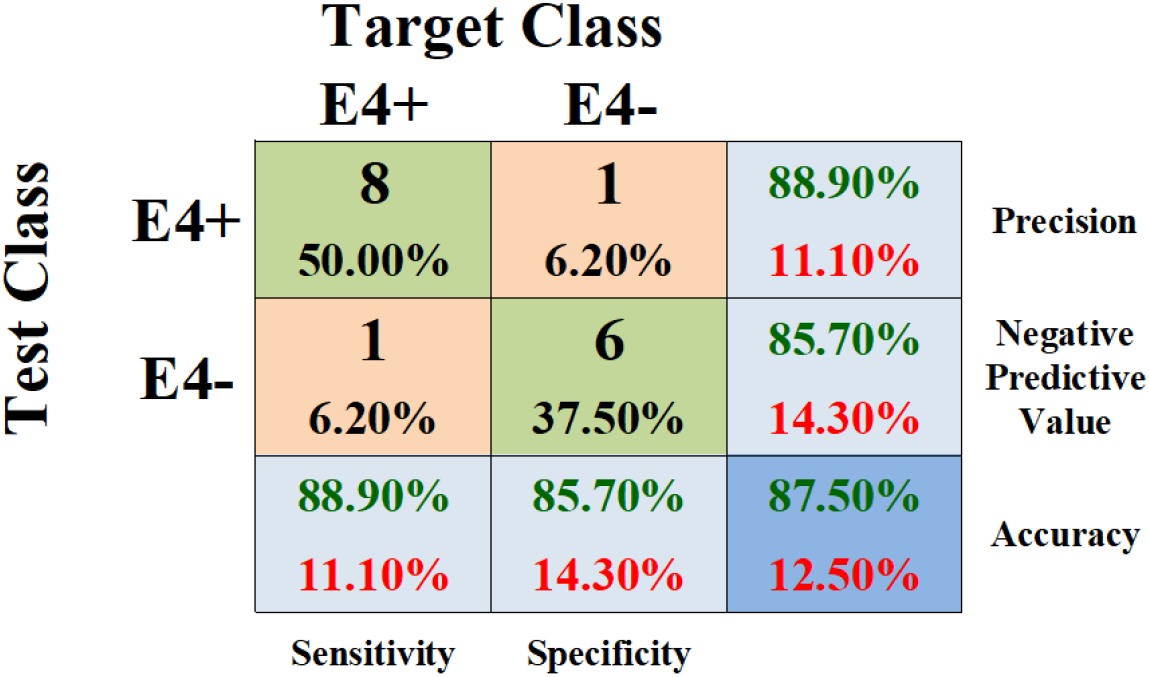
Confusion matrix for binary classification of APOE-ε4 carrier state (E4+ vs. E4−).

## Discussion

The present study investigated the impact of APOE-ε4 genotype on lip movement kinematics during a color-word interference task, focusing on how mild cognitive interference modulates speech motor control. Descriptive statistics revealed moderate effect sizes for several kinematic features despite non-significant p-values, supporting the interpretation that speech motor responses under cognitive load may reflect early neuromotor vulnerability in APOE-ε4 carriers. The classification results provide compelling preliminary evidence that speech kinematic responses to cognitive interference may serve as sensitive early indicators of APOE-ε4-associated risk for AD, even in cognitively normal older adults.

The classification model achieved a high level of accuracy (87.5%) in distinguishing between APOE-ε4 carriers and non-carriers using three kinematic speech features. The most discriminative feature combination included lip movement duration during the pre-interference segment, average lip movement speed during the interference segment, and the ΔDuring–Pre difference in lip movement range.

### Speech motor adjustments reveal cognitive-motor vulnerability in APOE-ε4 carriers

Altered articulatory movement duration observed during the pre-interference segment reflects anticipatory motor adjustments and resource allocation in preparation for upcoming cognitive interference. This has been attributed to proactive modulation of speech motor planning, where speakers change (i.e., typically slow down) articulatory movements to buffer against predicted instability and to facilitate error monitoring as cognitive demands increase.^57,58^ Predictive feedforward mechanisms within the motor system further contribute by prolonging movement duration to accommodate potential error margins under cognitive stress.^59^ Consistent with this interpretation, our descriptive data showed that pre-interference lip movement duration was moderately shorter in ε4+ individuals (d = −0.53), suggesting altered anticipatory motor strategies in this group. The inclusion of this feature in the final classification model underscores the potential diagnostic utility of pre-task articulatory dynamics in identifying APOE-ε4 status.

Similarly, prior research suggests that individuals under cognitive interference often adopt more conservative articulation strategies, such as slower and larger articulatory excursions, to maintain intelligibility and motor stability.^53,60,61^ In our study, the ΔDuring–Pre difference in movement range emerged as a key feature, suggesting that changes in articulatory flexibility under cognitive load are informative for group classification. Although the machine learning model does not directly quantify the direction of group-level changes, our descriptive analysis revealed a smaller absolute ΔDuring–Pre difference in lip movement range in the ε4+ group (0.205 mm^3^) compared to the ε4− group (0.609 mm^3^), with a moderate effect size (d = −0.371). This reduced dynamic modulation may indicate diminished neuromotor adaptability to cognitive demands in ε4+ individuals.

Altered articulatory speed during the interference condition of a cognitive-motor task has been linked to increased susceptibility to cognitive-motor conflict, reflecting difficulty in maintaining speech precision while resolving competing demands. Prior research suggests that when cognitive control resources are heavily taxed, neural resources are reallocated from motor execution to processes such as conflict monitoring and inhibition, resulting in slower and less precise articulatory movements.^62–65^ These effects are especially pronounced in at-risk populations, including older adults and individuals with executive dysfunction, who tend to exhibit greater slowing and variability in movement during cognitive interference,^66^ further highlighting the close interplay between executive function and speech motor control. ^62–65^ This pattern aligns with findings from dual-task gait studies, where APOE-ε4 carriers show slower walking speeds and greater reductions in step length when simultaneously engaged in executive function tasks.^67^ Although these findings focus on gross motor behavior, they illustrate a broader compensatory mechanism in which motor systems prioritize stability, often through slower or more deliberate movements, under increased cognitive load. In the present study, average lip movement speed during the interference segment did not differ significantly between groups (d = −0.18), but it contributed substantially to classification accuracy, reinforcing its relevance as a speech timing feature linked to cognitive load sensitivity.

### Clinical utility of speech-based biomarkers for APOE-ε4

These findings support the hypothesis that subtle speech motor differences in cognitively intact APOE-ε4 carriers can be detected through articulatory kinematics, particularly under mild cognitive load. A classification accuracy of 87.50%, along with precision and sensitivity values of 88.90% and specificity of 85.70%, demonstrates the effectiveness of speech-based features in identifying ε4+ individuals while minimizing false positives, an essential balance in clinical contexts. Unlike traditional neuropsychological assessments that focus on cognitive decline, speech kinematics offer a behaviorally grounded, low-burden, and non-invasive window into early neuromotor changes, especially those affecting articulatory timing and precision. These features show promise as early indicators of genetic risk and may augment existing multimodal approaches to stratification, screening, and individualized risk monitoring.

Their scalability, accessibility, and suitability for remote, longitudinal data collection make them especially valuable in underserved or resource-limited settings. Because they can be elicited passively and without extensive testing infrastructure, these features are uniquely suited to complement traditional clinical and biomarker-based assessments.

Importantly, our results suggest that APOE-ε4 carriers with intact cognition may already exhibit early motor abnormalities, reinforcing the idea that motor deficits can precede overt cognitive decline. This aligns with previous research showing that individuals with the APOE-ε4 allele experience accelerated motor decline independent of detectable cognitive impairment.^6,25^ The underlying mechanisms likely involve multiple neurobiological pathways. APOE-ε4 is strongly associated with increased amyloid-β deposition and tau pathology, which disrupt neural circuits critical for motor control.^68–70^ Additionally, synaptic loss and dysfunction in motor regions may impair the fine motor coordination necessary for speech production.^68,70^ Neuroinflammation and cerebrovascular dysfunction, both characteristic features of APOE-ε4 carriers, could further contribute to early motor deficits.^71–73^ While our findings provide indirect behavioral evidence of such disruptions, future work integrating speech kinematics with imaging and molecular markers will be necessary to clarify the mechanistic basis of these early articulatory changes.

## Conclusion

This study highlights distinct alterations in speech motor control among APOE-ε4 carriers, particularly under conditions of cognitive-linguistic interference during a color-word interference task. Machine learning analysis identified a combination of temporal and spatial speech kinematic features that accurately classified ε4+ individuals with 87.50% accuracy. These findings suggest that subtle disruptions in articulatory timing and flexibility may serve as early behavioral markers of APOE-ε4–associated risk for AD. The altered kinematic properties observed in ε4+ carriers may reflect early neural vulnerabilities related to motor adaptability, inhibitory control, and cognitive-motor integration.

Future research should investigate whether these motor signatures intensify over time and correlate with emerging cognitive decline, offering insight into the temporal evolution of neurodegenerative risk. Moreover, incorporating speech kinematic assessments into precision medicine frameworks may enhance personalized risk profiling, especially when combined with genetic and fluid biomarkers. These results support the broader utility of speech-based measures as scalable, non-invasive tools for early detection and longitudinal monitoring in at-risk populations.

## Study Limitations

Several limitations of this study should be acknowledged. First, the small sample size (N = 16) limits statistical power and reduces the ability to detect significant group differences or reliably assess the directionality of changes in individual kinematic features. Second, the cross-sectional design precludes evaluation of whether speech kinematic differences in APOE-ε4 carriers evolve over time or predict future cognitive decline. Third, the absence of neuroimaging or other biological markers restricts mechanistic interpretation, making it difficult to directly link observed motor changes to specific brain network alterations associated with APOE-ε4. Fourth, the lack of an MCI comparison group limits the ability to determine whether these articulatory changes are specific to preclinical AD risk or reflect more general patterns of cognitive aging. Finally, the unbalanced gender distribution within the ε4+ group limits generalizability and precludes analysis of potential sex differences, which are particularly relevant given the elevated AD risk in female APOE-ε4 carriers.^74–76^

## Consent Statement

Recruitment and consent procedures adhered to Mass General Brigham Healthcare (MGH) and HIPAA regulations. The study was approved by the MGB Institutional Review Board (IRB Protocol #2021P001460).

Before participating in testing, administrators thoroughly explained the experimental tasks and procedures, addressing any questions participants had. Relevant consent forms were provided, and each form was reviewed point by point with potential participants by a research staff member prior to obtaining their signature. Participants were explicitly informed of their right to withdraw from the study at any time during the experiment without any consequences and all participants provided written informed consent prior to participation.

## Data Availability

The raw speech recordings generated during the current study contain personally identifiable information and cannot be shared publicly for privacy and ethical reasons. A de-identified dataset comprising the extracted kinematic features that support the findings of this study is available from the corresponding author upon reasonable request.

## Author Contribution

M.D. contributed to data analysis, statistical methodology, and manuscript preparation. L.C.H. contributed to data analysis, statistical methodology, and manuscript preparation. J.R.G. contributed to the interpretation of findings, reviewed the manuscript, and provided feedback. B.D.R. supported data collection, contributed to IRB submission and approval, reviewed the manuscript, and provided feedback. A.T.C. analyzed the kinematic data. N.V.B. assisted with data collection, reviewed the manuscript, and provided feedback. D.H.S. contributed to participant characterization, manuscript review, and feedback. M.E. played a leading role in study conceptualization and design, data collection, data analysis, interpretation of findings, and manuscript writing. All authors contributed to the article and approved the submitted version.

